# The effect of non-pharmaceutical interventions on COVID-19 cases, deaths and demand for hospital services in the UK: a modelling study

**DOI:** 10.1101/2020.04.01.20049908

**Authors:** Nicholas G. Davies, Adam J. Kucharski, Rosalind M. Eggo, Amy Gimma, CMMID COVID-19 working group, W. John Edmunds

**Affiliations:** Department of Infectious Disease Epidemiology, London School of Hygiene & Tropical Medicine, Keppel Street, London, WC1E 7HT, UK

## Abstract

**Background:** Non-pharmaceutical interventions have been implemented to reduce transmission of SARS-CoV-2 in the UK. Projecting the size of an unmitigated epidemic and the potential effect of different control measures has been critical to support evidence-based policymaking during the early stages of the epidemic.

**Methods:** We used a stochastic age-structured transmission model to explore a range of intervention scenarios, including the introduction of school closures, social distancing, shielding of elderly groups, self-isolation of symptomatic cases, and extreme “lockdown”-type restrictions. We simulated different durations of interventions and triggers for introduction, as well as combinations of interventions. For each scenario, we projected estimated new cases over time, patients requiring inpatient and critical care (intensive care unit, ICU) treatment, and deaths.

**Findings:** We found that mitigation measures aimed at reducing transmission would likely have decreased the reproduction number, but not sufficiently to prevent ICU demand from exceeding NHS availability. To keep ICU bed demand below capacity in the model, more extreme restrictions were necessary. In a scenario where “lockdown”-type interventions were put in place to reduce transmission, these interventions would need to be in place for a large proportion of the coming year in order to prevent healthcare demand exceeding availability.

**Interpretation:** The characteristics of SARS-CoV-2 mean that extreme measures are likely required to bring the epidemic under control and to prevent very large numbers of deaths and an excess of demand on hospital beds, especially those in ICUs.

**Research in Context:** *Evidence before this study:* As countries have moved from early containment efforts to planning for the introduction of large-scale non-pharmaceutical interventions to control COVID-19 outbreaks, epidemic modelling studies have explored the potential for extensive social distancing measures to curb transmission. However, it remains unclear how different combinations of interventions, timings, and triggers for the introduction and lifting of control measures may affect the impact of the epidemic on health services, and what the range of uncertainty associated with these estimates would be.

*Added value of this study:* Using a stochastic, age-structured epidemic model, we explored how eight different intervention scenarios could influence the number of new cases and deaths, as well as intensive care beds required over the projected course of the epidemic. We also assessed the potential impact of local versus national targeting of interventions, reduction in leisure events, impact of increased childcare by grandparents, and timing of triggers for different control measures. We simulated multiple realisations for each scenario to reflect uncertainty in possible epidemic trajectories.

*Implications of all the available evidence:* Our results support early modelling findings, and subsequent empirical observations, that in the absence of control measures, a COVID-19 epidemic could quickly overwhelm a healthcare system. We found that even a combination of moderate interventions – such as school closures, shielding of older groups and self-isolation – would be unlikely to prevent an epidemic that would far exceed available ICU capacity in the UK. Intermittent periods of more intensive lockdown-type measures are predicted to be effective for preventing the healthcare system from being overwhelmed.

## Introduction

The novel coronavirus SARS-CoV-2 has spread to multiple countries after causing an initial outbreak of disease (COVID-19) in Wuhan, China [1]. Early evidence indicated SARS-CoV-2 was capable of sustained human-to-human transmission [2] and could cause severe disease [3], with a higher risk of severe and fatal outcomes in older individuals [4]. The first two cases of COVID-19 in the United Kingdom (UK) were confirmed on 31 January 2020. Although implementation of testing, isolation and contact tracing likely slowed early transmission [5], it was not sufficient to contain the outbreak in the UK.

Following the introduction of extensive control measures in Wuhan in late January, including—among other measures—travel restrictions, social distancing, and requirements for residents to stay within their homes, there was a substantial decline in local transmission [6–8]. Social distancing measures, such as closure of schools, bars, restaurants, and constraints on individual movements and interactions, are now in place in many countries with the aim of reducing transmission of SARS-CoV-2 [9,10]. However, it remains unclear precisely how the timing, duration, and intensity of different measures targeting transmission and burden can reduce the impact of COVID-19. Here, based upon scenarios originally presented to scientific advisory bodies in the UK, we use a mathematical model to assess the potential impact of different control measures for mitigating the burden of COVID-19, and evaluate possible medium-term scenarios as the most restrictive short-term measures are eventually lifted.

## Methods

### Dynamic transmission model

We analysed a stochastic compartmental model stratified into 5-year age bands, with individuals classified according to current disease status (**Fig. 1**) and transmission between groups based on UK social mixing patterns ([11,12]; full details in Supplementary Information). After infection with SARS-CoV-2 in the model, susceptible individuals pass through a latent period before becoming infectious, either with a preclinical and then clinical infection, or with a subclinical infection, before recovery or isolation. We refer to those infections causing few or no symptoms as subclinical. We assume older individuals are more likely to show clinical symptoms [11]. The model tracks 66.4 million people aggregated to the 186 county-level administrative units in England, Wales, Scotland, and Northern Ireland.

**Fig. 1.**
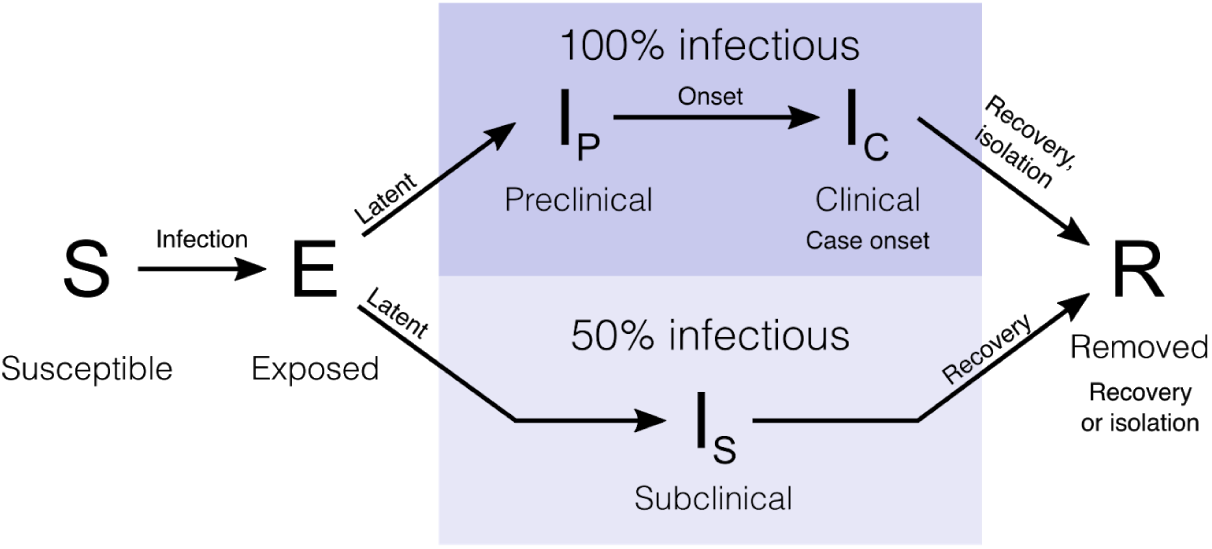
State transitions in the model. Individuals in the stochastic compartmental model are classified into susceptible, exposed, infectious (preclinical, clinical, or subclinical), and recovered states. The model is stratified into 5-year age bands and epidemics are simulated in the 186 county-level administrative units of the UK.

### Key model parameters

We collated multiple sources of evidence to estimate key model parameters (**Table S1**). In a meta-analysis, we estimated that the basic reproduction number, *R*_0_—which describes the average number of secondary infections caused by a typical primary infection in a completely susceptible population—was 2.7 (95% credible interval: 1.6–3.9) across settings without substantial control measures in place. We derived age-stratified case fatality ratios (CFR) to estimate a CFR that ranged substantially across age groups, from 0.1% in the 20–29 age group to 7.7% in the over-80 age group. Using these values along with the relationship between CFR and severe and critical cases, we also estimated the proportion of clinical cases in each age group that would require hospitalisation, which was 0.8% in the 20-29 age group and 62% in the over-80 age group (**Table S2**).

### Intervention scenarios

We explored a variety of non-pharmaceutical interventions, which we assumed would impact the rate of contact between individuals as well as the relative infectiousness of symptomatic individuals. Contact matrices were constructed by summing home, work, school, and “other” contacts calculated from survey data [12], with interventions altering the relative number of contacts of each type (**Table 2**). We simulated self-isolation of symptomatic individuals by decreasing their infectiousness by 35% during the intervention period. This was based on a calculation that approximately 70% of contacts occur outside the home; we assumed that these could be reduced by half for individuals under self-isolation, consistent with findings that accelerated case isolation in Shenzhen, China reduced transmission by 35% [13]. We included regular school closures for holidays in all models.

**Table 2.** Intervention scenarios. Each intervention was assumed to affect either a component of the contact matrix or the infectiousness of symptomatic individuals, reducing it to the percentage shown.

|  | Home contacts | Work contacts | School contacts | Other contacts | Infectiousness of symptomatic individuals |
| --- | --- | --- | --- | --- | --- |
| Baseline | 100% | 100% | 100% | 100% | 100% |
| School Closures | 100% | 100% | 0% | 100% | 100% |
| Social Distancing | 100% | 50% | 100% | 50% | 100% |
| Elderly Shielding | 100% | 25% (70+);<br>100% (others) | 100% | 25% (70+);<br>100% (others) | 100% |
| Self-Isolation | 100% | 100% | 100% | 100% | 65% |
| Combined | 100% | 25% (70+);<br>50% (others) | 0% | 25% (70+);<br>50% (others) | 65% |
| Intensive interventions<br>(see Supporting Information) | 100% | 25% (70+);<br>65% (others) | 100% (open)<br>0% (closed) | 16% (70+);<br>59% (others) | 65% |
| Lockdown | 100% | 10% | 10% (open);<br>0% (closed) | 10% | 65% |

### Statement on data availability

All analysis code and data are available at https://github.com/cmmid/covid-uk.

## Results

### Projections for an unmitigated epidemic

Simulations of an unmitigated COVID-19 epidemic resulted in a median 24 million (95% prediction interval: 16–30 million) clinical cases in the UK up to December 2021 (**Fig. 2; Table 4**). Under this scenario, 85% of the population (68–96%) would be infected by SARS-CoV-2, with roughly 40% of those infected showing clinical symptoms. In turn, this would result in a projected 370 thousand (250–470 thousand) deaths directly attributable to COVID-19, without accounting for any potential increase in the case fatality ratio caused by exceeding hospital capacity. The projected peak number of ICU beds required was 220 thousand (120–360 thousand). This is roughly 25–80 times ICU capacity in the UK, which we tallied at 4,562 beds [14–17] in the absence of any efforts to further expand capacity.

**Table 4.** Projected impact of 12-week interventions in the UK. Median and 95% prediction interval reported. Totals are calculated up to December 31, 2021.

|  | Base | School Closures | Social Distancing | Elderly Shielding | Self-Isolation | Combination |
| --- | --- | --- | --- | --- | --- | --- |
| **Total cases** | 24 M (16 M–30 M) | 18 M (11 M–25 M) | 17 M (10 M–23 M) | 17 M (10 M–23 M) | 18 M (10 M–24 M) | 18 M (11 M–24 M) |
| **Total deaths** | 370 k (250 k–470 k) | 280 k (150 k–400 k) | 250 k (140 k–360 k) | 220 k (130 k–330 k) | 260 k (140 k–390 k) | 280 k (150 k–390 k) |
| **Cases in peak week** | 4.2 M (2.5 M–6.4 M) | 2.8 M (940 k–6.2 M) | 2.2 M (1 M–4.2 M) | 2.9 M (1.1 M–6.4 M) | 2.7 M (900 k–6.1 M) | 3.5 M (1.2 M–5.3 M) |
| **Deaths in peak week** | 62 k (35 k–98 k) | 41 k (13 k–96 k) | 31 k (14 k–62 k) | 34 k (12 k–79 k) | 39 k (12 k–92 k) | 51 k (17 k–82 k) |
| **Peak ICU beds required** | 220 k (120 k–360 k) | 150 k (45 k–350 k) | 110 k (49 k–220 k) | 120 k (42 k–300 k) | 140 k (44 k–330 k) | 190 k (61 k–300 k) |
| **Peak non-ICU beds required** | 420 k (230 k–670 k) | 280 k (85 k–670 k) | 210 k (92 k–420 k) | 230 k (79 k–560 k) | 270 k (83 k–630 k) | 350 k (120 k–570 k) |
| **Time to peak cases (weeks)** | 11 (8.2–16) | 14 (10–25) | 19 (11–28) | 14 (9.2–21) | 14 (10–25) | 22 (16–36) |

**Fig. 2.**
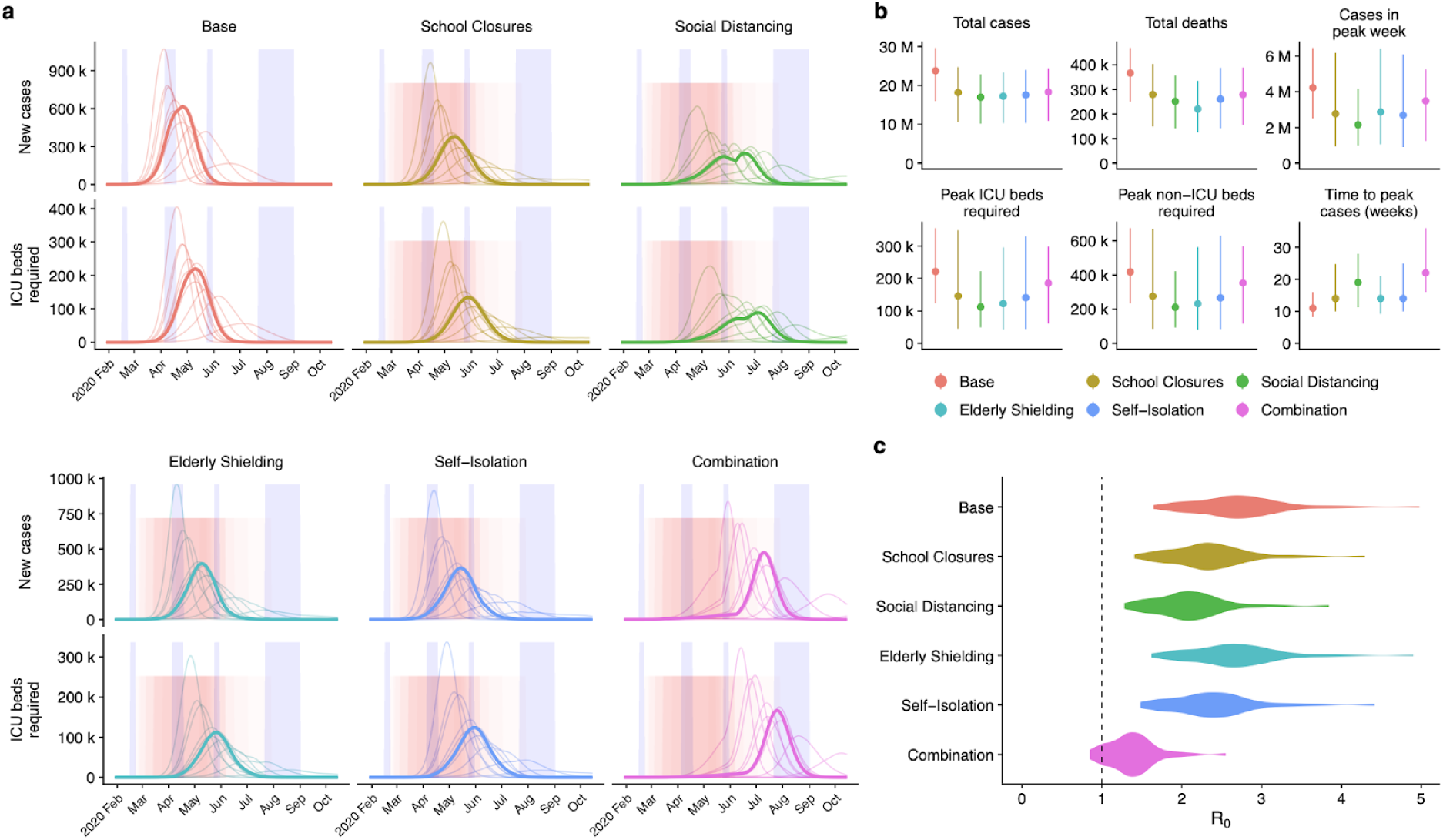
Impact of interventions lasting 12 weeks. (a) Daily new cases and ICU beds required over the course of the simulated scenarios in the UK. From 50 realisations of each projection, we show 11 representative simulations, corresponding to each decile of the total number of cases; the bold curve shows the simulation resulting in the median projected number of cases. Blue shaded regions show regular school closures, while the pink shaded region shows the distribution of 12-week interventions. (b) Summary of simulated outputs, in total number of clinical cases and deaths, clinical cases in peak week, peak ICU beds required, peak non-ICU beds required, and the time from seeding until peak of the epidemic. (c) The sampled distribution of the basic reproduction number, *R*_*0*_, under each intervention scenario.

### Impact of non-pharmaceutical interventions

Non-pharmaceutical interventions against previous epidemics—particularly school closures in response to pandemic influenza or SARS—have typically been put in place for periods of one week to three months [18]. Accordingly, we first evaluated a number of scenarios under which non-pharmaceutical interventions would be deployed for 12 weeks. The interventions we analysed were school closures; social distancing; shielding of the elderly; self-isolation of symptomatic individuals; and combinations of these policies (**Table 2**). These mitigation measures decreased the total number of cases by 70–75% and delayed the peak of the epidemic by 3–8 weeks on average (**Fig. 2a, b**). While social distancing was predicted to have the greatest impact on the total number of cases, elderly shielding was predicted to have the greatest impact on the number of deaths (**Table 4**).

We found that, when implemented alone, none of these shorter-duration interventions were able to decrease the healthcare need to below available capacity. We estimated that neither school closures, social distancing, elderly shielding, or self-isolation alone would reduce *R*_0_ enough to lead to a substantial decline in the total number of cases (**Fig. 2c**). In particular, school closures had a limited impact in our projections, despite our model accounting for substantial asymptomatic transmission among children. This contrasts with strategies aimed at suppressing the spread of pandemic influenza, for which school closures are often a key intervention [19].

Next, we sought to evaluate the potential impact of combining control measures. The most comprehensive of these involves deploying all four individual strategies at the same time. This combination strategy was projected to have a more marked impact on *R*_0_ (**Fig. 2c**), and in a small proportion (8%) of simulations, was sufficient to halt the epidemic altogether during the intervention period. However, lifting the interventions leads to a rapid resurgence of cases in the model, even when *R*_0_ had been kept below 1 during the intervention period.

### Triggering of interventions

When interventions have a limited duration, their impact can be influenced by timing. If interventions are triggered at the same time across all locations, they may arrive too early in some locations and too late in others. We therefore estimated the impact of triggering interventions at different times, both nationally and at a local level. We projected that triggering interventions locally instead of nationally could modestly reduce the total number of cases and deaths, as well as reduce peak demands on the healthcare system (**Fig. 3a, b; Table S3**). However, our simulations do not account for any differences in the implementation of or adherence to control measures which might arise from these timings varying in different parts of the country. Examining the simulated dynamics at a county level (**Fig. 3c**) shows that the timing of local epidemics may vary among counties, and highlights that epidemics at a local level are predicted to peak more sharply than they do across the entire UK.

**Fig. 3.**
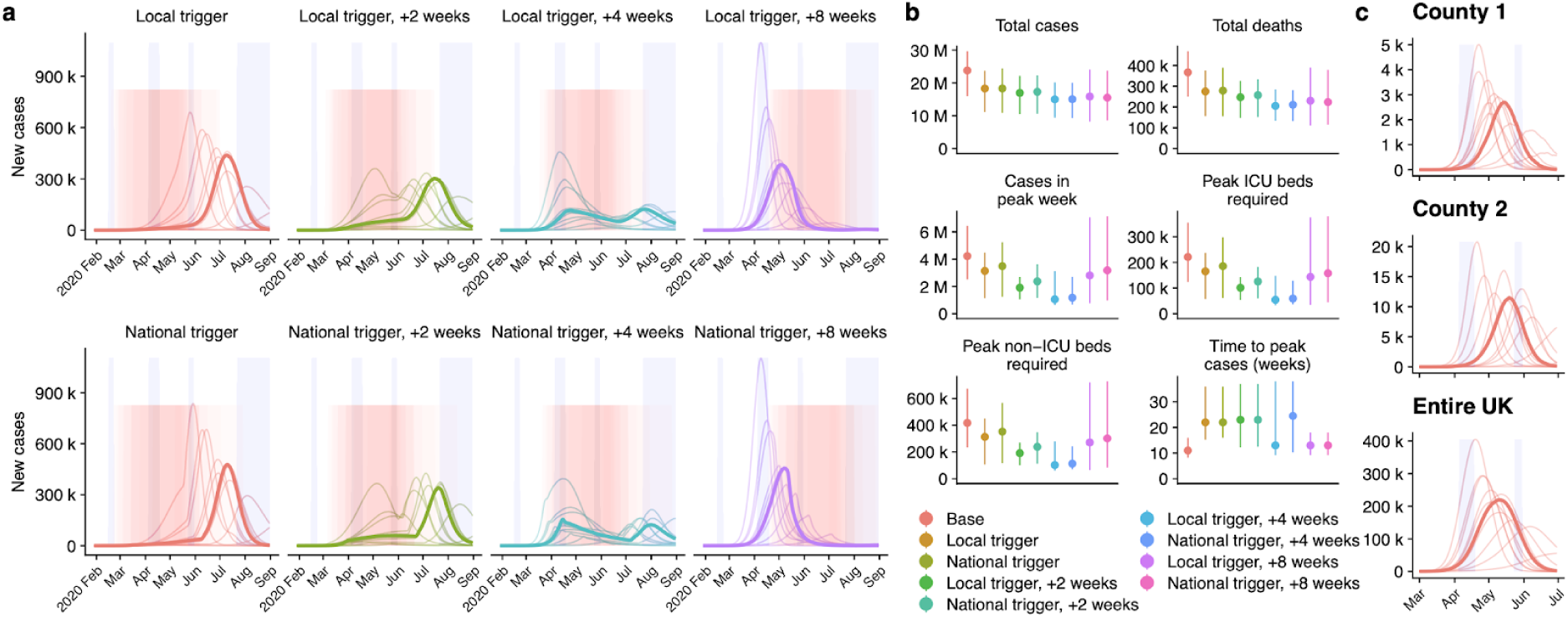
Local versus national triggering and timing of interventions. (a) Dynamics of the epidemic under local versus national triggers for introduction of interventions (pink shaded regions). Blue shaded regions show regular school closures, while the pink shaded region shows the intervention period. Bolded lines show daily incidence of cases in the median simulation under each scenario. (b) Summary of simulated outputs, in total number of clinical cases, deaths, clinical cases in peak week, peak ICU beds required, peak non-ICU beds required, and the time from seeding until peak of the epidemic (c) Illustration of peak timings of new cases varying across two counties in the UK, in comparison with predicted national trends, for a single simulation with no control interventions. Case incidence at a local scale is expected to rise and fall more rapidly than case incidence across the country as a whole.

Our projections also showed that, when only a short intervention is deployed, rather than centring measures over the peak (as predicted in the absence of control measures), it was preferable to trigger the intervention later in order to minimise the total health burden (**Fig. 3b**). This is because the introduction of control measures will change the timing of the peak relative to the baseline scenario (**Fig. 3a**). In particular, the most effective timing for introduction of measures could involve a delay of as much as four weeks (**Fig. 3b**). However, optimally timing an intervention may be more difficult in practice than these scenarios suggest, since here they are run with complete knowledge of when the simulated peak would occur in the absence of any intervention.

We concluded that a period of intense restrictions on interpersonal contacts, combined with shielding the most vulnerable members of society, had the potential to substantially reduce the burden of COVID-19 for as long as they were in place—but that this strategy alone, particularly if enacted only over relatively short timeframes, would not substantially reduce the overall impact of the COVID-19 epidemic.

### Leisure activities and older-adult care of children

As other countries in Europe began restricting mass gatherings, there was a question about the impact such measures might have in the UK, with a particular focus on stopping spectator sports [20]. By analysing the total attendance at spectator sports in the UK, we performed additional simulations to evaluate the potential impact of such restrictions (**Fig. 4a, Table S2**). Although yearly attendance at sporting events is high (75.1 million spectators per year [21]), even if we assume that people make the equivalent of their mean daily physical contacts at such events (i.e. 5 contacts per person, to make a total of 375 million), this number is very low relative to the number of yearly contacts which occur outside the context of sporting events (269 billion [12]). We estimated that stopping spectator sports would have little direct effect on the number of cases (**Fig. 4a, Table S4**). We simulated a more general reduction in leisure contacts—which mainly occur in pubs, bars, restaurants and cinemas—by reducing leisure contacts by 75%, and found a larger (though still modest) impact on the epidemic. Previous work on pandemic influenza has estimated that many individuals are likely to choose to avoid such settings, as they perceive them to be risky [22].

**Fig. 4.**
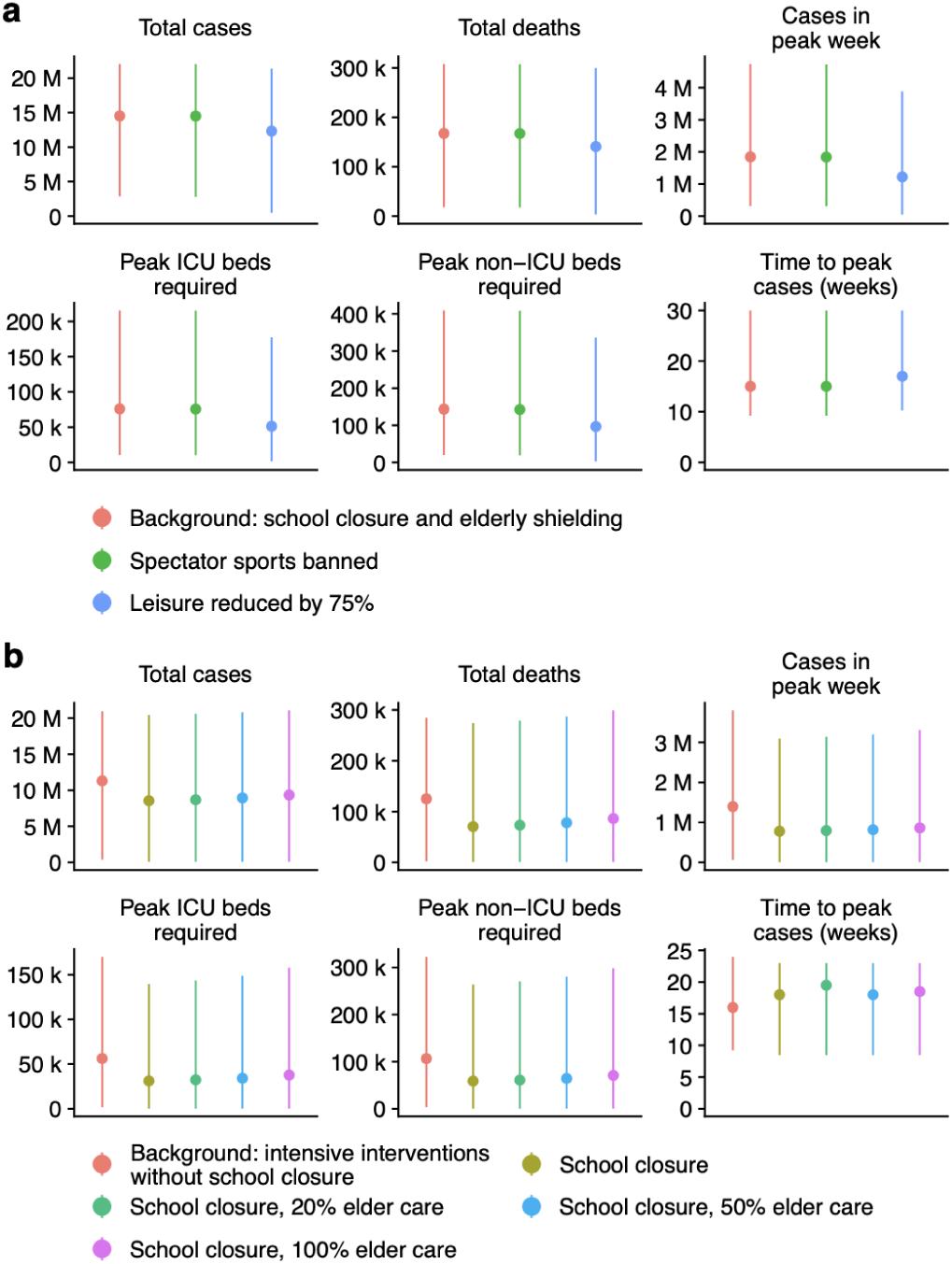
Impact of reducing leisure events and impact of increased childcare from older age groups. (a) Effect of banning spectator sports, and decreasing leisure activities on the total cases, total deaths, and peak number of cases, ICU beds, non-ICU beds, and the time to peak week in the simulated epidemics. The “Background” to which these interventions are compared is school closures plus elderly shielding. (b) Effect of varying increases in contacts between children and older adults during school closures and effect on the total cases, total deaths, and peak number of cases, ICU beds, non-ICU beds, and the time to peak week in the simulated epidemics. The “Background” to which these interventions are compared is the “Intensive Interventions” of Table 2, without any additional period of school closure.

We also evaluated the potential impact of schoolchildren being cared for by grandparents during weekdays as a result of school closures, because of concerns over whether this might counteract the benefit of closing schools as a result of higher-risk older adults being exposed to more transmission from children. We found that, over a period of school closure from 17th March to 1st September, one additional contact per weekday between children under 15 and an older individual (at least 55 years older than the child) could, in the worst case (i.e. high *R*_0_), almost entirely eliminate the benefit of closing schools in terms of the number of deaths and peak ICU bed occupancy over this period (**Fig. 4b, Table S5**).

### Intensive interventions and lockdown

As well as single 12-week measures, during the first half of March 2020 we also analysed the impact of longer-term and repeated interventions. On March 16th 2020, our group was advised that, supported by the results of modelling analyses from multiple sources (including our preliminary projections), a package of intensive interventions would be put in place, including a significant programme of social distancing, with a particular impact on leisure activities; workers being asked to work from home where possible; shielding of both elderly (70+) individuals and people in high-risk-groups of all ages; school closures; and self-isolation of symptomatic individuals. With these more concrete proposals, we updated our model to estimate the likely impact of the proposed strategy. We projected that the intensive interventions being proposed had the potential to delay the peak of the epidemic by 8 weeks on average (95% prediction interval: 1–50 weeks), and to reduce the total number of deaths by half (**Fig. 5a; Table 5**).

**Fig. 5.**
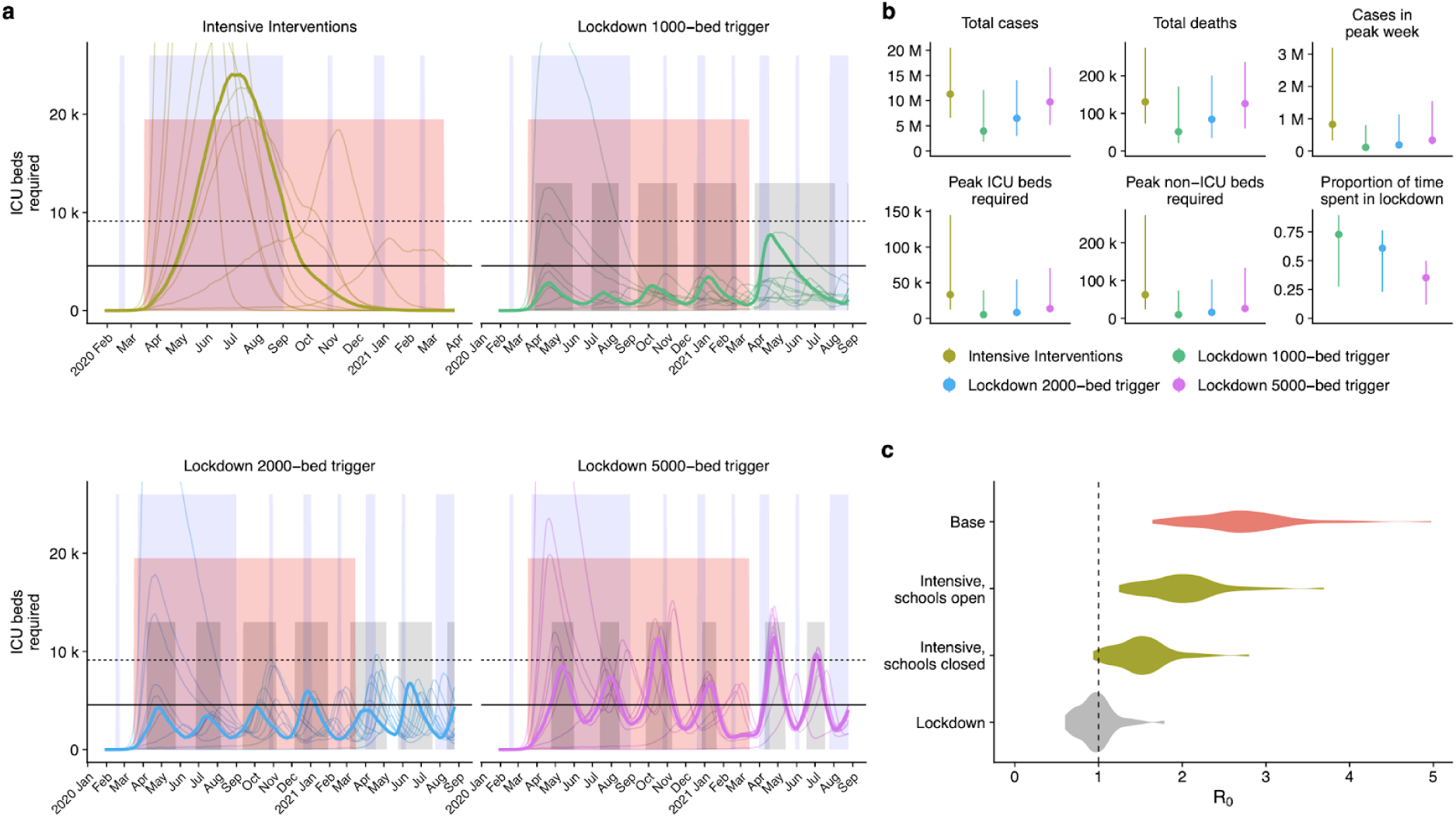
Projected impact of intensive control measures with reactive lockdowns. (a) Dynamics of the epidemic under different triggers for introduction and lifting of lockdowns (median timing of lockdowns shown as grey shaded areas). Bolded lines show ICU bed occupancy in the median run under each scenario. Horizontal guides show the estimated number of ICU beds in the UK as of January 2020 (solid line) and with a hypothetical doubling of capacity (dashed line). Blue shaded regions show school closures, while the pink shaded region shows a background period of intensive interventions. (b) Summary of epidemic runs. (c) Estimated distribution of *R*_0_ under three different interventions: intensive social distancing with schools open and closed, and lockdown.

**Table 5.** Projected impact of intensive control measures and lockdown in the United Kingdom. Median and 95% prediction interval reported. Totals are calculated up to December 31, 2021.

| statistic | Intensive Interventions | Lockdown with 1000-bed trigger | Lockdown with 2000-bed trigger | Lockdown with 5000-bed trigger |
| --- | --- | --- | --- | --- |
| Total cases* | 11 M (6.6 M–21 M) | 4 M (1.8 M–12 M) | 6.5 M (3 M–14 M) | 9.7 M (5.2 M–17 M) |
| Total deaths* | 130 k (73 k–270 k) | 51 k (21 k–170 k) | 84 k (34 k–200 k) | 130 k (60 k–240 k) |
| Cases in peak week | 820 k (330 k–3.2 M) | 110 k (79 k–800 k) | 190 k (110 k–1.1 M) | 330 k (200 k–1.5 M) |
| Deaths in peak week | 9.3 k (3.5 k–40 k) | 1.4 k (850–11 k) | 2.3 k (1.3 k–15 k) | 3.7 k (2.3 k–20 k) |
| Peak ICU beds required | 33 k (12 k–140 k) | 5 k (3.2 k–39 k) | 8.1 k (4.8 k–55 k) | 13 k (8.4 k–71 k) |
| Peak non-ICU beds required | 62 k (23 k–270 k) | 9.4 k (6.2 k–73 k) | 16 k (9 k–100 k) | 26 k (16 k–130 k) |
| Time to peak cases (weeks) | 19 (9.2–66) | 60 (8–96) | 46 (8–71) | 34 (8–63) |
| Proportion of time spent in lockdown (29 Jan 2020–31 Dec 2021) | — | 0.73 (0.27–0.9) | 0.61 (0.23–0.77) | 0.35 (0.12–0.5) |
| Total infected* | 28 M (18 M–48 M) | 11 M (4.3 M–33 M) | 18 M (6.9 M–36 M) | 27 M (12 M–41 M) |

\* Simulations were run to December 31, 2021, so total cases, deaths, and infections under the lockdown strategies may not reflect the full number during the entire epidemic.

In spite of this substantial reduction in burden, the projections still showed a large number of cases (6.6–21 million), and a large number of ICU beds (12–140 thousand) occupied during the peak of the epidemic (**Fig 5b**). Indeed, we projected that ICU bed capacity could be exceeded by 5-fold or more for several weeks. While we could not explicitly predict the impact of this on mortality rates, this would likely lead to an increased case fatality ratio.

We had previously presented scenarios on 11th March showing that shorter, repeated periods of particularly strict restrictions on movement—”lockdowns”—could be used to supplement a longer-term, more moderate package of interventions, with lockdowns to be deployed as needed to prevent the resources of the health system becoming overburdened. Accordingly, we supplemented the intensive interventions with lockdowns phased in when ICU bed capacity reached certain thresholds, which would be kept in place until ICU bed usage fell back below the same trigger threshold, to then be brought in again as needed.

We found that adding these periods of lockdown would still result in a high number of ICU beds being occupied, but at much lower levels than the scenario without lockdowns (**Fig. 5a**). Lockdown periods were sufficient to bring *R*_0_ near or below 1 (**Fig. 5c**), and hence to lead to a reduction in total COVID-19 cases (**Fig. 5b**). We found that, depending on the threshold ICU bed occupancy at which lockdown periods were triggered, there was a tradeoff between having fewer, longer lockdown periods (lower threshold) and having more, shorter lockdown periods (higher threshold), with the higher thresholds resulting in less time spent in lockdown overall, but higher peak demands on ICU bed capacity (**Table 5**). Lower thresholds also resulted in more individuals remaining susceptible at the end of the simulation period, potentially increasing the total duration for which recurrent lockdowns would need to be maintained. We assumed that lockdowns would be triggered at a national level rather than at a local level, and that the trigger threshold would not change over time. There are likely to be better strategies for selecting timing and duration of lockdowns. However, we presented our results as supporting evidence that periodic lockdowns could reduce the burden of COVID-19 without measures being in place indefinitely.

## Discussion

Using an age-structured transmission dynamic model, we explored different scenarios for COVID-19 transmission and control in the UK. We found that moderate interventions lasting for 12 weeks, such as school closures, self-isolation or shielding of elderly groups, would likely not have been sufficient to control the epidemic and to avoid far exceeding available ICU capacity, even when these measures were used in combination. However, we estimated that a scenario in which more intense lockdown measures were implemented for shorter periods may be able to keep projected case numbers at a level that would not overwhelm the health system.

The model presented here is subject to several limitations. Because the model does not explicitly structure individuals by household, we are unable to evaluate the impact of measures based on household contacts, e.g. household quarantine, i.e., where all members of a household with a suspected COVID-19 case remain in isolation. Such contact-targeted measures could increase the impact of a package of interventions by limiting spread in the community. However, the presence of asymptomatic infections [23] means that isolation based on symptomatic case identification would be unlikely to fully prevent ongoing transmission. We also do not include individual level variation in transmission (i.e. ‘superspreading events’, [24]). There are several examples of such events for COVID-19 [25], and individual-level variation is likely important in influencing the success of control measures in the very early stages of an outbreak [5]. However, as outbreaks of directly-transmitted infections become larger, the population-level dynamics will predominantly be driven by the average mixing pattern between key epidemiological groups, particularly between different ages [11,26]. We therefore used a stochastic model implementation to capture variation in these population-level dynamics. We also assumed that subclinically-infected individuals were 50% as infectious as clinical cases. A study of 2,147 close contacts in Ningbo, China estimated that the mean onward transmission from asymptomatic infections was 65% (95% HDI: 20–120%) that of symptomatic cases [23]. However, symptomatic cases were found to be more likely to generate new symptomatic infections compared to asymptomatic infections. This suggests that the overall relative contribution of asymptomatic individuals to new infections may be lower than 65%, and hence 50% is a plausible assumption. We used mixing matrices for the UK measured in 2006 [12], and changes in contact patterns since then may alter the potential effect of interventions. The fractions of hospitalisation, ICU use, and death are estimated using data from China, and any differences in UK populations could affect our estimates of health care demand.

The results we present here summarize the key analyses and scenarios we presented to decision makers during February and March 2020, which evolved continuously as additional information became available. A reasonable worst-case scenario with and without school closures, focusing on Birmingham as an illustrative example, was presented to the Scientific Pandemic Influenza Group on Modelling (SPI-M), which gives expert advice to the UK Department of Health and Social Care and wider UK government, on 26th February 2020. This was followed by an exploration of national-level impact of shorter-duration interventions (as in **Fig. 2**) presented on 2nd March 2020, which explored various assumptions concerning intervention length and efficacy. We expanded our analysis to explicitly cover all counties in England and analysed the timing of measures, and local versus national deployment of interventions (as in **Fig. 3**), on 8th March 2020. Our analyses of the impact of curtailing sporting events and leisure activities (as in **Fig. 4a**), and of the potential impact of repeated lockdown measures (as in **Fig. 5**), were presented on 11th March 2020. Our sensitivity analysis for increased child-grandparent contacts (as in **Fig. 4b**) was presented on 17th March 2020. The results shown in the main text are based on the final version of the model, and reflect our current state of knowledge about the transmission dynamics of COVID-19. However, our overall conclusions about the relative effectiveness of different strategies for reducing the burden of COVID-19 in the UK are the same as those presented to decision makers in real-time.

## Data Availability

All analysis code and data are available at https://github.com/cmmid/covid-uk.

## Acknowledgements

We thank Anna Foss, Quentin Leclerc, Ruwan Ratnayake, and David Simons for comments on a draft manuscript.

## Supplementary Information

### Dynamic transmission model

We analyse a stochastic compartmental model (**Fig. 1**) stratified into 5-year age bands, with time approximated in discrete 6-hour steps. The model tracks 66.4 million UK residents aggregated to the 186 county-level administrative units across England, Wales, Scotland, and Northern Ireland. We run 50 stochastic realizations for each projection.

We assume that the population initially consists of susceptible individuals (S), who become exposed (E) after effective contact with an infectious person. After an incubation period lasting 4 days on average, exposed individuals of age *i* will develop either a clinical infection with probability *y*_*i*_, or a subclinical infection with probability 1 − *y*_*i*_. Clinical cases begin with a preclinical but infectious (I_P_) state lasting 1.5 days on average; these individuals then progress to a clinically infected state (I_C_), which we assume marks the onset of a clinical case. We assume that subclinical infections (I_S_) are half as transmissible as preclinical and clinical infections. Regardless of whether they are clinically or subclinically infected, individuals remain infectious for 5 days on average and are then removed (R) from the infectious state; we assume that removed individuals are immune to reinfection over the 1–2 years over which we simulated the epidemic. Hospitalisations and deaths from COVID-19 are assumed to occur among clinical cases only, and we assume that the clinical outcome of a case does not impact upon transmission dynamics.

The amount of time a given individual spends in states *E, I*_*P*_, *I*_*C*_, or *I*_*S*_ is drawn from distributions *d*_*E*_, *d*_*P*_, *d*_*C*_, or *d*_*S*_, respectively (**Table S1**). The force of infection for an individual in age group *i* at time *t*,, where t is defined in 6 hour time steps.

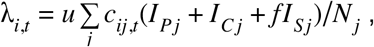

is the rate at which susceptible individuals enter the exposed state. Here, *u* is an individual’s susceptibility to infection upon contact with an infectious person, *c*_*ij,t*_ is the number of age-*j* individuals contacted by an age-*i* individual per day at time *t, f* is the relative infectiousness of a subclinical case, and 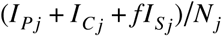 is the effective probability that a random age-*j* individual is infectious.

To calculate the basic reproductive number, *R*_0_, defined as the average number of secondary infections generated by a typical infectious individual in a fully susceptible population, we define the next generation matrix as

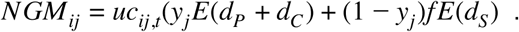

Then, *R*_0_ is the absolute value of the dominant eigenvalue of the next generation matrix.

### Key parameters of the transmission model

We used a serial interval of 6.5 days based on published studies [1,27,28], and assumed that the length of the preclinical period was 30% of the total period of clinical infectiousness [13]. From this, we fixed the mean of the latent period to 4 days, the mean duration of preclinical infectiousness to 1.5 days, and the mean duration of clinical infectiousness to 3.5 days.

The basic reproduction number *R*_0_ was estimated by synthesizing the results of a literature review (**Fig. S1**). For each reported value of the basic reproduction number, we matched a flexible PERT distribution (a shifted beta distribution parameterised by minimum, maximum, and mode) to the median and confidence interval reported in each study. We sampled from the resulting distributions, weighting each study equally, to obtain estimates of *R*_0_ for our simulations.

The age-specific clinical fraction *y*_*i*_ was adopted from an estimate based on case data from 6 countries [11], and the relative infectiousness of subclinical cases, *f*, was assumed to be 50% relative to clinical cases, as we assumed in a previous study [11].

We used the Office for National Statistics data on the population by age for each of 186 county-level subdivisions of the UK [29], comprising non-metropolitan counties, metropolitan counties, unitary authorities and London boroughs in England; unitary authorities in Wales; council areas in Scotland; and local government districts in Northern Ireland (hereafter referred to as “counties”). We used contact data from the POLYMOD study [12] and the R package socialmixr [30] to generate age-stratified contact matrices for the UK, generating separate contact matrices for each county according to the population structure for that county, assuming that the number of age-*j* contacts made by an age-*i* individual scales with the number of age-*j* individuals in a county.

We assumed that epidemics in each county are seeded by 2 individuals per day for 28 days; after this point, further seeding has very little impact owing to extensive community transmission. Seeding times are staggered so that London boroughs are seeded on a random day in the first week of the simulation, and other locations are seeded on a random day in the first four weeks of the simulation. We assume that transmission between counties is negligible, instead allowing the staggered seeding of infection among counties to simulate the process of gradual introduction of the epidemic across the UK. The start date of the model is 29th January 2020, which we chose by visually aligning model-predicted deaths to the daily number of COVID-19 deaths reported in the UK [31] up to 27th March.

### Hospital burden estimation

To calculate ICU and non-ICU beds in use through time, we scaled age-stratified symptomatic cases by age-specific hospitalisation and critical outcome probability, then summed to get the total number of hospitalised and critical cases. We then distributed hospitalised cases over time based on expected time of hospitalisation and duration admitted. We assumed gamma-distributed delays, with the shape parameter set equal to the mean, for: delay from symptom onset to hospitalisation of mean 7 days (standard deviation 2.65) [32,33]; delay from hospitalisation to discharge / death for non-ICU patients of mean 8 days (s.d. 2.83) [34]; delay from hospitalisation to discharge / death for ICU patients of mean 10 days (s.d. 3.16) [33]; and delay from onset to death of mean 22 days (s.d. 4.69) [32,33]. We calculated the age-specific case fatality ratio based on data from the COVID-19 outbreak in China and on the Diamond Princess cruise ship. We first calculated the naive case fatality ratio, nCFR, (i.e. deaths/cases) for each age group, then scaled down the naive CFR based on a correction factor estimated from data from the Diamond Princess [35] to give an adjusted CFR. We then calculated risk of hospitalisation based on the ratio of severe and critical cases to cases (18.5%) and deaths to cases (2.3%) in the early China data, which we took to imply 8.04 times more hospitalisations than deaths in each age group. We assumed all age groups had a 30% risk of requiring critical care if hospitalised [33].

### Derivation of contact rates for the “Intensive Interventions” scenario

For the “Intensive Interventions” scenario, we assumed that 30% of workers would be able to work from home [36], reducing work and transport contacts (11% of “other” contacts) among the low-risk general population (assumed to be 90% of adults under 70) by 30%. We also assumed leisure contacts (45% of “other” contacts) would decrease by 75% in this population. We assumed that work and “other” contacts would be reduced by 75% among the high-risk general population (10% of under-70s) through shielding. Among those aged 70 and above, we assumed that 75% of work and other contacts would be reduced through shielding; we then further reduced transport contacts by 30% and leisure contacts by 75%.

### R_0_ meta-analysis

We sampled R_0_ from a consensus distribution (**Fig. S1**) derived from published sources available at the time projections were made. We sampled across all studies, with each study weighted equally. The distribution overall has a mean *R*_0_ of 2.68, with a standard deviation of 0.57. We used a normal distribution with these parameters for our simulations.

## Supplementary Figures

**Fig. S1.**
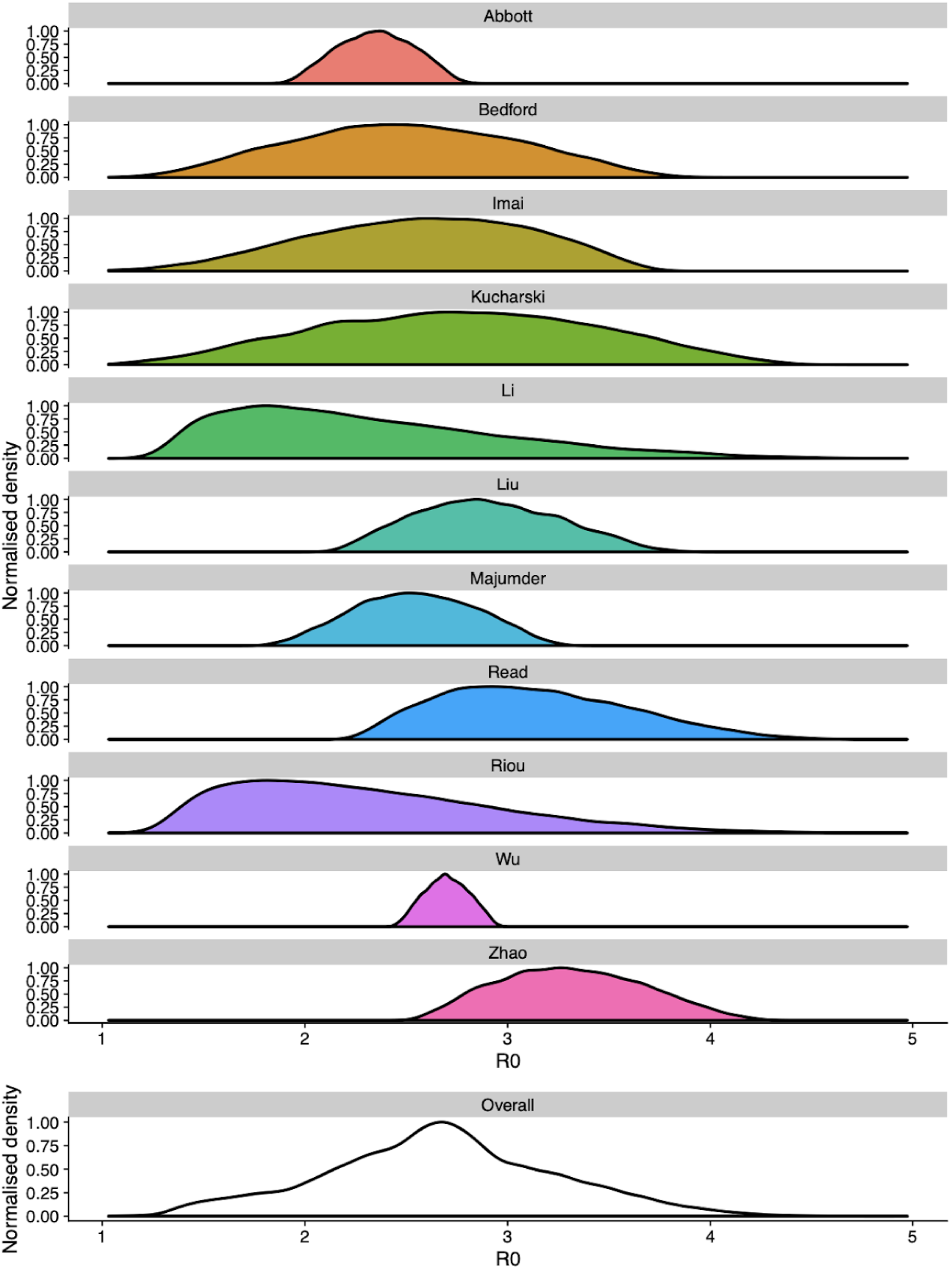
R_0_ distribution used. Sources used can be found among refs. [1,6,37–45].

## Supplementary Tables

**Table S1.** Model parameters.

| Parameter | Description | Value | Reference |
| --- | --- | --- | --- |
| $d_E$ | Latent period (E to $I_p$ and E to $I_s$; days) | $\sim \text{gamma}(\mu = 4.0, k = 4)$ | [1,27,28] |
| $d_P$ | Duration of preclinical infectiousness ($I_p$ to $I_C$; days) | $\sim \text{gamma}(\mu = 1.5, k = 4)$ | [13] |
| $d_C$ | Duration of clinical infectiousness ($I_C$ to R; days) | $\sim \text{gamma}(\mu = 3.5, k = 4)$ | [1,27,28] |
| $d_S$ | Duration of subclinical infectiousness ($I_s$ to R; days) | $\sim \text{gamma}(\mu = 5.0, k = 4)$ | Assumed to be the same duration as total infectious period for clinical cases, including preclinical transmission |
| | Incubation period (E to $I_C$; days) | $d_E + d_P$ ; mean 5.5 days | Derived |
| | Serial interval (days) | $d_E + (y_i(d_P + d_C) + (1 - y_i)d_S)/2$ ; mean 6.5 days | Derived |
| $u$ | Susceptibility to infection on contact | Calculated from $R_0$ | Derived |
| $y_i$ | Probability of clinical symptoms on infection for age group $i$ | Estimated from case distributions across 6 countries | [11] |
| $f$ | Relative infectiousness of subclinical cases | 50% | Assumed |
| $c_{ij}$ | Number of age-$j$ individuals contacted by an age-$i$ individual per day | UK-specific contact matrix | [12] |
| $N_i$ | Number of age-$i$ individuals | Demographic data | [29] |
| $\Delta t$ | Time step for discrete-time simulation | 0.25 days | |
| | Delay from onset to hospitalisation (days) | $\sim \text{gamma}(\mu = 7, k = 7)$ | [32,33] |
| | Duration of hospitalisation in non-ICU bed (days) | $\sim \text{gamma}(\mu = 8, k = 8)$ | Duration based on NHS data for J12: viral pneumonia, not elsewhere classified [34]. |
| | Duration of hospitalisation in ICU bed (days) | $\sim \text{gamma}(\mu = 10, k = 10)$ | [33] |
|  | Proportion of hospitalised cases that require critical care | 30% | [33] |
| | Delay from onset to death (days) | $\sim \text{gamma}(\mu = 22, k = 22)$ | [32,33] |

**Table S2.** Age-specific hospitalisation and fatality risk. Based on estimates from the early COVID-19 outbreak in China [4].

| Age group | Cases (China) | Deaths (China) | Pop. (%; China) | Naïve CFR | Adjusted CFR | Hospitalised |
| --- | --- | --- | --- | --- | --- | --- |
| 0–9 | 416 | 0 | 12.0% | 0.0% | 0.00% | 0.0% |
| 1–10 | 549 | 1 | 11.6% | 0.2% | 0.09% | 0.8% |
| 20–29 | 3619 | 7 | 13.5% | 0.2% | 0.10% | 0.8% |
| 30–39 | 7600 | 18 | 15.6% | 0.2% | 0.12% | 1.0% |
| 40–49 | 8571 | 38 | 15.6% | 0.4% | 0.23% | 1.9% |
| 50–59 | 10008 | 130 | 15.0% | 1.3% | 0.68% | 5.4% |
| 60–69 | 8583 | 309 | 10.4% | 3.6% | 1.87% | 15.1% |
| 70–79 | 3918 | 312 | 4.7% | 8.0% | 4.14% | 33.3% |
| 80–89 | 1408 | 208 | 1.8% | 14.8% | 7.68% | 61.8% |

**Table S3.** Projected impact of control measures in the United Kingdom depending upon local versus national triggering and according to shift from centring on predicted peak. Median and 95% prediction interval reported. Totals are calculated up to December 31, 2021.

|  | Base | Local trigger | National trigger | Local trigger, +2 weeks | National trigger, +2 weeks | Local trigger, +4 weeks | National trigger, +4 weeks | Local trigger, +8 weeks | National trigger, +8 weeks |
| --- | --- | --- | --- | --- | --- | --- | --- | --- | --- |
| **Total cases** | 24 M (16 M–30 M) | 18 M (11 M–24 M) | 18 M (11 M–24 M) | 17 M (10 M–22 M) | 17 M (11 M–22 M) | 15 M (9.4 M–20 M) | 15 M (9.3 M–20 M) | 16 M (8.2 M–24 M) | 16 M (8.6 M–24 M) |
| **Total deaths** | 370 k (250 k–470 k) | 280 k (160 k–370 k) | 280 k (150 k–390 k) | 250 k (150 k–330 k) | 260 k (150 k–330 k) | 210 k (130 k–290 k) | 210 k (130 k–280 k) | 230 k (110 k–390 k) | 220 k (120 k–380 k) |
| **Cases in peak week** | 4.2 M (2.5 M–6.4 M) | 3.1 M (1.1 M–4.5 M) | 3.5 M (1.2 M–5.3 M) | 1.9 M (1 M–2.7 M) | 2.4 M (1.2 M–3.6 M) | 1.1 M (630 k–3.1 M) | 1.2 M (670 k–2.7 M) | 2.8 M (760 k–7.1 M) | 3.2 M (970 k–7.1 M) |
| **Deaths in peak week** | 62 k (35 k–98 k) | 45 k (16 k–66 k) | 51 k (17 k–82 k) | 28 k (15 k–40 k) | 35 k (17 k–50 k) | 15 k (9 k–41 k) | 17 k (10 k–36 k) | 40 k (9.5 k–99 k) | 43 k (12 k–100 k) |
| **Peak ICU beds required** | 220 k (120 k–360 k) | 170 k (57 k–240 k) | 190 k (61 k–300 k) | 100 k (53 k–140 k) | 130 k (59 k–180 k) | 54 k (32 k–150 k) | 59 k (37 k–130 k) | 140 k (34 k–380 k) | 160 k (44 k–380 k) |
| **Peak non-ICU beds required** | 420 k (230 k–670 k) | 310 k (110 k–450 k) | 350 k (120 k–570 k) | 190 k (100 k–270 k) | 240 k (110 k–350 k) | 100 k (61 k–280 k) | 110 k (70 k–240 k) | 270 k (64 k–720 k) | 300 k (84 k–730 k) |
| **Time to peak cases (weeks)** | 11 (8.2–16) | 22 (15–36) | 22 (16–36) | 23 (12–37) | 23 (12–37) | 13 (9.2–38) | 24 (10–38) | 13 (9.2–18) | 13 (9.2–18) |

**Table S4.**
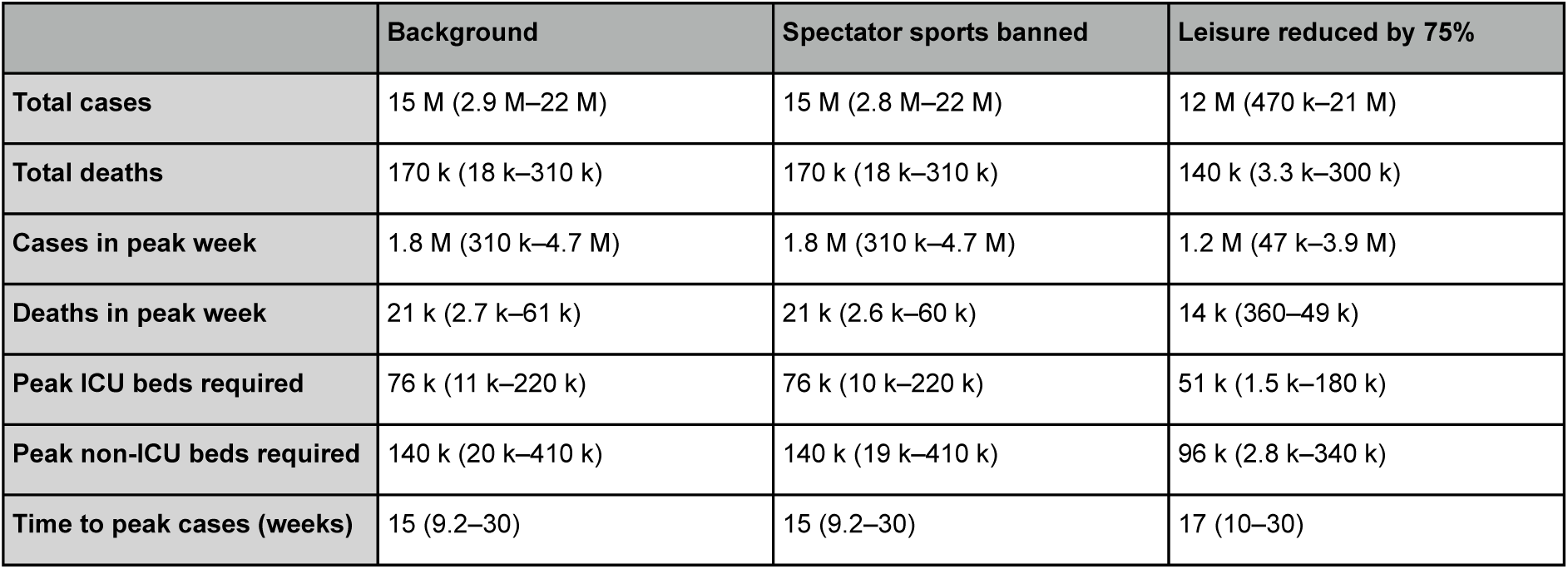
Projected impact of control measures relating to leisure activities in the United Kingdom. Median and 95% prediction interval reported. Totals are calculated up to September 1st, 2020.

**Table S5.** Projected impact of school closures, depending upon additional contact between children and the elderly, in the United Kingdom. Median and 95% prediction interval given. Totals are calculated up to July 20th, 2020. “Care by elders” denotes the percentage of children under 15 for which one additional daily contact with an individual 55 years older or more is added to simulations during school closures.

| statistic | Background | School closure | School closure, 20% care by elders | School closure, 50% care by elders | School closure, 100% care by elders |
| --- | --- | --- | --- | --- | --- |
| Total cases | 11 M (400 k–21 M) | 8.5 M (64 k–20 M) | 8.7 M (66 k–21 M) | 8.9 M (63 k–21 M) | 9.3 M (75 k–21 M) |
| Total deaths | 130 k (2 k–280 k) | 71 k (550–270 k) | 73 k (590–280 k) | 78 k (530–290 k) | 86 k (650–300 k) |
| Cases in peak week | 1.4 M (63 k–3.8 M) | 780 k (3.5 k–3.1 M) | 790 k (3.6 k–3.1 M) | 820 k (3.4 k–3.2 M) | 860 k (4.5 k–3.3 M) |
| Deaths in peak week | 16 k (320–48 k) | 8.8 k (40–39 k) | 9.1 k (49–40 k) | 9.6 k (42–42 k) | 11 k (54–44 k) |
| Peak ICU beds required | 56 k (1.7 k–170 k) | 31 k (140–140 k) | 32 k (140–140 k) | 34 k (150–150 k) | 38 k (180–160 k) |
| Peak non-ICU beds required | 110 k (3.4 k–320 k) | 59 k (260–260 k) | 61 k (250–270 k) | 64 k (260–280 k) | 71 k (330–300 k) |
| Time to peak cases (weeks) | 16 (9.2–24) | 18 (8.5–23) | 20 (8.5–23) | 18 (8.5–23) | 18 (8.5–23) |

